# Prospective Genomic Surveillance of Severe Febrile Illness in Tanzanian Children Identifies High Mortality and Resistance to First-Line Antibiotics in Bloodstream Infections

**DOI:** 10.1101/2025.05.25.25328306

**Authors:** Teresa B. Kortz, Victoria T. Chu, Raya Y. Mussa, Erasto M. Sunzula, Victoria M. Mlele, Cecilia L. Msemwa, Lushona K. Mathias, Namala P. Mkopi, Ryan J. Ward, Kathleen S. Sun, Jazmin M. Baez Maidana, Anneka M. Hooft, Juma A. Mfinanga, Joseph L. DeRisi, Hendry R. Sawe, Joel P. Manyahi, Charles R. Langelier

## Abstract

We evaluated the prevalence, pathogen profile, and antimicrobial resistance (AMR) patterns of bloodstream infections (BSIs) among 392 children with severe febrile illness who presented (July 26, 2022-September 20, 2023) to a referral hospital in Tanzania. We identified a causative pathogen in 9.8% (n=38) of participants. Blood culture analysis confirmed BSI in 5.2% (n=20) of participants with a case fatality rate of 45%. Whole genome sequencing (WGS) of blood culture isolates identified gram-negative bacteria (*Escherichia coli, Klebsiella pneumoniae*) as the predominant pathogens, many exhibiting extended-spectrum beta-lactamase (ESBL) resistance genes (CTX-M-15, CTX-M-27), rendering them resistant to first-line antimicrobials. We also observed probable nosocomial transmission in ventilated patients based on phylogenetic analyses of tracheal aspirate isolates. There is an urgent need for enhanced AMR surveillance, empiric antibiotic regimens tailored to local AMR patterns, culture-independent diagnostics, and robust infection control practices in resource-limited settings to mitigate BSI-related mortality and minimize nosocomial transmission risk.

**Funding:** NIAID K23AI144029 (TBK), NIAID K23AI185326 (VTC), Chan Zuckerberg Biohub (JLD, CRL)

**Article Summary Line:** Genomic surveillance of severe febrile illness in Tanzanian children reveals high mortality rates and widespread resistance to first-line antibiotics, highlighting the urgent need for tailored treatments and enhanced antimicrobial resistance monitoring.

## Introduction

Severe febrile illness (SFI) is a major cause of childhood death in Sub-Saharan Africa (SSA).(1–3) Among children hospitalized for SFI, bloodstream infections (BSIs) carry particularly high mortality,(4–7) with treatment increasingly complicated by antimicrobial resistance (AMR).(8, 9) Despite global efforts and international guidelines mandating empiric antimicrobial treatment, mortality from SFI has not improved over the past decade.(10, 11)

AMR is a major global public health threat, with an estimated 4.71 million deaths associated with resistant bacterial infections in 2021.(12) AMR is agnostic to country, region, and income level. Despite this, poverty, health inequity, and the absence of regulatory oversight contribute to the underlying causes and consequences of AMR.(13) A recent Global Burden of Disease study reported that SSA accounted for 20% of the global burden of AMR-associated deaths, the highest of any region.(12) Furthermore, an estimated 59% of AMR-attributable deaths occurred in children under five years of age in SSA as compared to 0.18% in a high-income region.(12) The prevalence of AMR in children hospitalized for SFI in SSA, however, is largely unknown.

Reducing the burden of AMR in SSA is hindered by a lack of data on prevalence, phenotypic resistance patterns, and genomic features of resistant microbes, especially among children. For many countries, it is unknown whether empiric antibiotic therapy for pediatric SFI is appropriately matched to pathogens causing BSI. Moreover, recent studies demonstrate a concerning and alarming increase in resistance to several first-line antimicrobials, emphasizing the urgent need to more deeply characterize AMR patterns.(4, 14–16)

Even less is known about the genomic features of pathogens responsible for community-acquired BSI in the pediatric population and the antimicrobial resistance genes (ARGs) they harbor. To address these gaps, we carried out a prospective cohort study in Tanzanian children with SFI and assessed the prevalence and clinical features of BSI. Using whole genome sequencing (WGS), we identified causative microbial pathogens and their ARGs, including associations with mortality and subsequent nosocomial respiratory infections.

## Materials and Methods

### Study Design and Participants

We screened all children presenting to the Emergency Medicine Department (EMD) at Muhimbili National Hospital (MNH), the national referral hospital for Tanzania in Dar es Salaam, from July 26, 2022-September 20, 2023. The EMD is the presenting department for all acutely ill and injured children and is a 24-hour, full-capacity, government-funded EMD, caring for approximately 60,000 patients annually. Children comprise approximately 25% of the patient volume, with approximately 75% of those presenting subsequently admitted.

Inclusion criteria were children aged ≥28 days to ≤14 years with SFI and a guardian fluent in Kiswahili or English. SFI was defined as fever (axillary temperature >38°C or history of fever) associated with this illness with ≥1 warning sign(s): respiratory distress, altered mental status, impaired perfusion, or at-risk for dehydration.(17) Respiratory distress was defined as tachypnea for age, increased work of breathing (presence of retractions, flaring, or head bobbing), or hypoxia (O_2_ saturation <92%).(18) Altered mental status was defined as ‘verbal’ or less on the Alert-Verbal-Pain-Unresponsive (AVPU) scale,(19) or history of convulsion within the last 24 hours in a patient without known epilepsy.(20) Impaired perfusion was the presence of ≥1 sign(s): hypotension for age, cold extremities, or severe tachycardia for age.(18, 21) At-risk for dehydration was defined as the presence of ≥1 of: ≥5 loose stools over the past 24 hours; ≥3 loose stools over the past 24 hours and emesis; ≥3 emeses over the past 24 hours; decreased urine output per guardian report; or inability to drink or breastfeed.(20, 22)

We excluded children who presented in cardiac arrest, weighed <4 kg, had acute trauma or burns, had a fever not thought to be due to an infection (e.g., rheumatologic disease), had known epilepsy with the sole warning sign of altered mental status due to convulsion, and those whose guardians were not available or declined to participate. MNH cares for a disproportionate number of children with malignancy and congenital heart disease; children known to have either of these conditions were excluded to prevent a biased sample and so that the study population would be more representative of a general pediatric population. The primary outcome of interest was in-hospital mortality; the secondary outcome was prevalence of BSI, which required a positive blood culture for likely bacterial or fungal pathogen. The sample size was determined *a priori* to detect an absolute difference in mortality of 20% between participants with vs. without BSI, assuming a BSI rate of 6%. We followed the STROBE reporting guidelines (**Supplemental Table 1**).(23)

The study was approved by the following ethics committees: MUHAS (DA.282/298/01.C/374), the Tanzanian National Institute for Medical Research (NIMRI) (NIMR/HQ/R.8a/Vol. IX/3576), and the University of California, San Francisco (19–27627). We extracted data from the hospital’s electronic and paper medical record systems and from the guardian. We obtained written informed consent from all participants’ guardians and verbal participant assent when appropriate. Participants received routine standard of care, including intravenous antibiotics, intravenous fluids, and antimalarial medications as needed. Children living with HIV were continued on their antiretrovirals. Permission to publish was obtained from NIMRI.

### Laboratory Procedures

Blood was collected from participants via venipuncture and using sterile technique within four hours of presentation. Point-of-care testing in the EMD included malaria screening (SD bioline-Pf), HIV screening (HIV-1/2 Alere for all participants, confirmatory testing for positive tests), blood sugar, and hemoglobin. For a blood culture, 1.5-3.0 mL of blood were collected in BD BACTEC™ Peds Plus/F culture vials (Becton Dickinson, BD). In a subgroup of participants requiring intubation with suspected respiratory infection, a tracheal aspirate was collected using sterile mucus extractors. All samples were immediately transported to the MUHAS Bacteriological Research Laboratory at room temperature for processing.

Blood culturing was performed using a BD BACTEC FX40 (Becton Dickinson, BD) instrument according to the manufacturer’s instructions. Positive cultures underwent gram-stain and sub-culturing onto each of the following plates: sheep blood agar (BA), chocolate agar (CA), and MacConkey agar (MAC). BA and CA plates were incubated at 35°C with 5-10% carbon dioxide (CO_2_) and MAC plates were incubated at 37°C; all plates were incubated for 18-24 hours.

For tracheal aspirates, we inoculated 1µl of the aspirate onto BA, CA, and MAC plates. BA and CA plates were incubated at 37°C in 5% CO_2_, MAC plates were incubated aerobically at 37°C, and all plates were incubated for 18-24 hours. We performed identification and susceptibility testing on isolates with bacterial growth of ≥10^5^ colony-forming units (CFU)/mL.

We used API20E (BioMérieux, Marcy, I’Etoile, France) and Staphaurex (Remel Europe Ltd, Dartford, UK) instruments for the identification of *Enterobacterales* and *Staphylococcus aureus,* respectively. Antimicrobial susceptibility was determined by the Kirby Bauer disk diffusion method following international standards.(24) For *Enterobacterales* that were resistant to ceftazidime and/or ceftriaxone, we confirmed extended-spectrum β-lactamase (ESBL) production by double disk approximation methods. Quality control and results were confirmed by the study clinical microbiologist (JM). The following organisms were considered blood contaminants and excluded from analysis: coagulase-negative *Staphylococcus* spp., *Corynebacterium* spp., *Micrococcus* spp., *Bacillus* spp., and *Brevibacterium* spp. *Candida* spp. were considered contaminants in tracheal aspirates. Contaminants were identified biochemically and genomically.

For WGS, DNA was extracted from cultures using the Zymo Quick-DNA Fungal/Bacterial Miniprep kit (Zymo Research). A 20ng sample of total nucleic acid from each sample was enzymatically sheared with fragmentase (New England Biolabs, Ipswich, MA USA) and libraries were constructed for sequencing using the NEBNext Ultra II Library Prep Kit (New England Biolabs). Samples were adaptor-ligated and multiplexed with unique dual-indexing primers, PCR amplified for 15 cycles, and SPRI cleaned to retain fragments with a median size of 450 base pairs. Libraries were quantified by Tapestation 4200 (Agilent, Santa Clara, CA USA), pooled, and underwent paired-end 150 base-pair sequencing on an Illlumina NextSeq 550.

Following demultiplexing, we processed raw sequencing files through the open-source CZ ID pipeline (https://czid.org/) to detect microbial species and strain (Illumina mNGS pipeline v8.3) and antimicrobial resistance genes (ARGs) (AMR pipeline v1.3.2).(25) CZ AMR pipeline implements the Comprehensive Antibiotic Resistance Database (CARD) Resistance Gene Identifier (RGI) tool,(26, 27) which aligns quality-controlled reads against a continually updated and curated reference database of ARG sequences. We included ARGs specific to the following antibiotic classes: aminoglycoside, beta-lactam, chloramphenicol, diaminopyrimidine/sulfonamide, fluoroquinolone, fosfomycin, macrolide-lincosamide-streptogramin (MLS), nitroimidazole, peptide, and tetracycline. Only ARGs with 100% read coverage breadth were retained for downstream analyses. We identified ESBL genes detected among *Enterobacterales* family culture isolates.

We evaluated genetic relatedness of sequenced isolates using the SNP Pipeline for Infectious Disease (SPID).(28, 29) SPID utilizes minimap2 to align short reads to a primary reference genome and call a consensus nucleotide at each genome position, thus enabling the calculation of a single nucleotide polymorphism (SNP) distance between all pairs of samples.(30) Maximum likelihood phylogenetic analyses were then performed using RAxML-NG following Multiple Sequence Alignment (MSA) methods and phylogenetic trees constructed using best tree fit.(31) Isolates with <10 SNPs across >80% of the pathogen genome were considered to represent probable transmission events.

### Clinical Data

Data were extracted from medical charts and included patient characteristics, medical interventions, diagnostic results, hospital course, and final hospital outcomes. Severity of illness was characterized using the Lambaréné Organ Dysfunction (LOD) Score, a simple mortality prediction score.(32) Research staff entered data into a secure, electronic REDCap database hosted and maintained locally on the secure MNH server using a study tablet.(33) Enrolled participants were evaluated from the time of presentation to the EMD until hospital discharge.

### Statistical Analysis

We used univariate descriptive statistics to summarize cohort-level information. We tested for associations between participant characteristics and in-hospital mortality using t-tests or Wilcoxon rank-sum tests (continuous variables) and χ^2^ or Fisher’s exact tests (categorical variables). A p-value <0.05 was considered statistically significant. We fit a logistic regression model to explore the association between the presence of a BSI and mortality and adjusted for confounding factors based on their empirical significance in the literature (age,(2, 34) HIV infection,(2) malnutrition,(35) immunization status(2)). We performed statistical analyses using Stata/MP 18.0 (StataCorp, TX), and generated figures using Prism for Mac v10.0 (GraphPad, https://www.graphpad.com) and RStudio (v2024.09.0+375).

## Results

We screened 8983 children who presented to MNH; 720 were deemed eligible and 392 were enrolled (**Figure 1**). Median cohort age was 17.8 months (IQR 9.1, 42.7) and 57.7% (n=226) were male (**Table 1**). Among all participants, 30.8% (n=121) had moderate-severe malnutrition, 1.8% (n=7) were living with HIV, 4.6% (n=18) tested positive for malaria, and 24.5% (n=95) required pediatric intensive care unit (PICU) admission.

**Figure 1.**
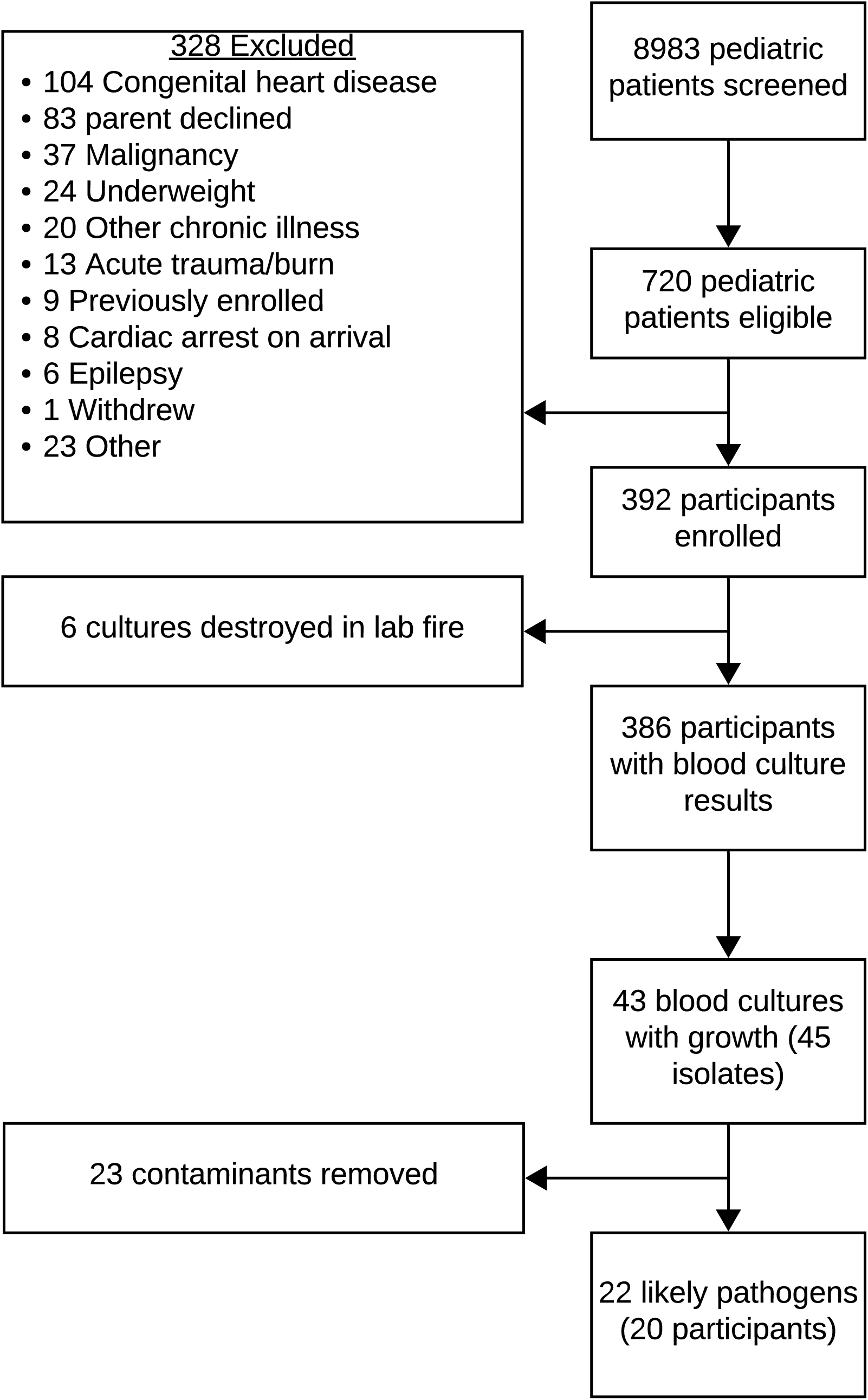
Flowchart depicting participant enrollment and blood culture isolates available for whole genome sequencing in Tanzanian children with severe febrile illness.

**Table 1.** Participant characteristics for children with severe febrile illness in Tanzania by survival status.

| Baseline Characteristic | Total (n=392) | Survivors (n=323) | Non-survivors (n=69) | p value |
| --- | --- | --- | --- | --- |
| Age (months), median (IQR) | 17.8 (9.1, 42.7) | 19.9 (10.3, 46.7) | 11.4 (6.6, 23.9) | 0.002 |
| Male sex, n (%) | 226 (57.7) | 187 (57.9) | 39 (56.5) | 0.83 |
| Moderate-Severe malnutrition, n (%) | 121 (30.9) | 87 (26.9) | 34 (49.3) | <0.001 |
| HIV positive, n (%) | 7 (1.8) | 2 (0.6) | 5 (7.2) | <0.001 |
| Malaria positive, n (%) | 18 (4.6) | 15 (4.6) | 3 (4.3) | 0.92 |
| Fully vaccinated, n (%) | 389 (99.2) | 321 (99.4) | 68 (98.6) | 0.47 |
| LOD score, mean (SD) | 1.1 (0.9) | 0.9 (0.8) | 2.1 (0.8) | <0.001 |
| Hemoglobin g/dL, median (IQR) | 9.6 (8.1, 11.1) | 9.9 (8.5, 11.2) | 8.6 (6.9, 10.1) | <0.001 |
| White blood cell count per uL, median (IQR) | 10.3 (6.9, 15.1) | 9.9 (6.6, 14.2) | 13.9 (9.6, 19.0) | <0.001 |
| Pre-Hospital antibiotics, n (%) | 181 (46.2) | 134 (41.5) | 47 (68.1) | <0.001 |
| Blood stream infection, n (%) | 20 (5.2) | 11 (3.4) | 9 (13.4) | <0.001 |
| **Outcome** |  |  |  |  |
| PICU admission, n (%) | 95 (24.5) | 43 (13.5) | 52 (75.4) | <0.001 |
| Intubation, n (%) | 62 (15.8) | 21 (6.5) | 41 (59.4) | <0.001 |

HIV, Human Immunodeficiency Virus; LOD, Lambaréné Organ Dysfunction; IQR, Interquartile Range; RDT, Rapid Diagnostic Test; SD, Standard Deviation; uL, microliter; g, gram; dL, deciliter; PICU, Pediatric Intensive Care Unit

All-cause in-hospital mortality was 17.6% (n=69). The prevalence of culture-confirmed BSI among those with available blood cultures (n=386) was 5.2% (n=20) and the case fatality rate was 45% (n=9/20). Mortality was significantly associated with BSI (9 [13.4%] vs. 11 [3.4%], p-value<0.001), mean LOD score (2.1 (SD 0.8) vs. 0.9 (SD 0.8), p-value<0.001), and PICU admission (52 [75.4%] vs. 43 [13.5%], p-value<0.001). The unadjusted and adjusted odds ratios for mortality in participants with BSI compared with those without was 4.4 [95% CI 1.7, 11.0] and 3.9 [95% CI 1.5, 10.1], respectively. Compared with surviving participants, those who died were significantly younger (11.4 months [IQR 6.6, 23.9] vs. 19.9 months [10.3, 46.7], p-value=0.002), had increased moderate-severe malnutrition (34 [49.3%] vs. 87 [26.9%], p-value<0.001), and were more likely to be living with HIV (5 [7.2%] vs. 2 [0.6%], p-value<0.001).

Prior to EMD presentation, 46.2% (n=181) of participants received antibiotics, among whom 45.3% (n=82/181) received ceftriaxone (**Table 2**). Empiric antibiotics were administered in the EMD to 63.3% (n=248) participants, and it was primarily ceftriaxone (91.9%, n=228/248) and metronidazole (40.3%, n=100/248). The most common in-hospital antibiotics included ceftriaxone (72.5%, n=177/244), metronidazole (13.1% n=32/244), and amoxicillin-clavulanate (13.1%, n=32/244). The most common infectious diagnoses were acute watery diarrhea (26%, n=101), sepsis (13.9%, n=54), and pneumonia (11.8%, n=46), while 20.8% (n=81) of participants did not have an infection diagnosed (**Supplemental Table 2**).

**Table 2.**
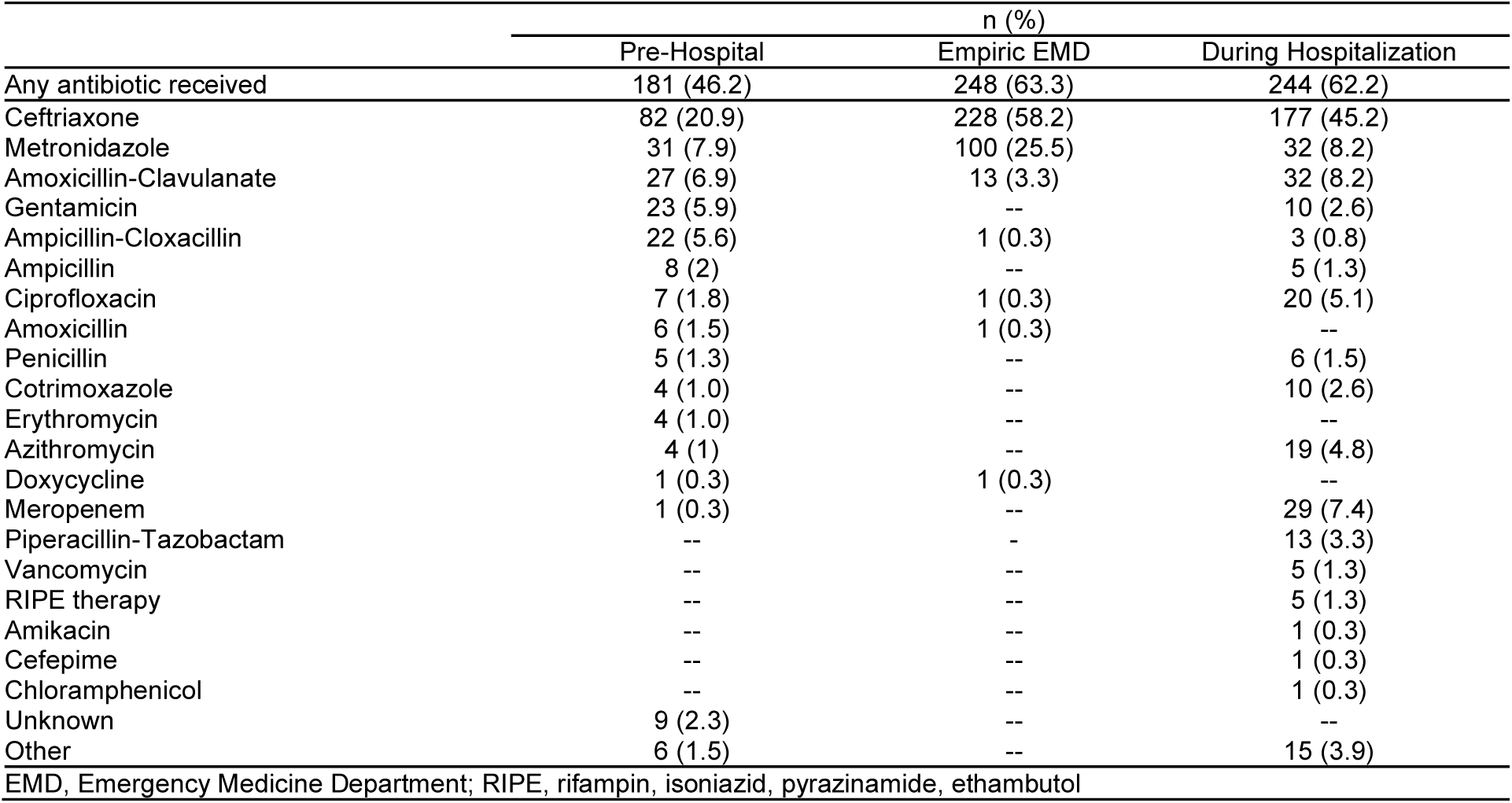
Antibiotic administration pre-hospital and empiric antibiotic administration in the Emergency Medicine Department for children with severe febrile illness in Tanzania (n=392).

|  | n (%) |  |  |
| --- | --- | --- | --- |
|  | Pre-Hospital | Empiric EMD | During Hospitalization |
| Any antibiotic received | 181 (46.2) | 248 (63.3) | 244 (62.2) |
| Ceftriaxone | 82 (20.9) | 228 (58.2) | 177 (45.2) |
| Metronidazole | 31 (7.9) | 100 (25.5) | 32 (8.2) |
| Amoxicillin-Clavulanate | 27 (6.9) | 13 (3.3) | 32 (8.2) |
| Gentamicin | 23 (5.9) | -- | 10 (2.6) |
| Ampicillin-Cloxacillin | 22 (5.6) | 1 (0.3) | 3 (0.8) |
| Ampicillin | 8 (2) | -- | 5 (1.3) |
| Ciprofloxacin | 7 (1.8) | 1 (0.3) | 20 (5.1) |
| Amoxicillin | 6 (1.5) | 1 (0.3) | -- |
| Penicillin | 5 (1.3) | -- | 6 (1.5) |
| Cotrimoxazole | 4 (1.0) | -- | 10 (2.6) |
| Erythromycin | 4 (1.0) | -- | -- |
| Azithromycin | 4 (1) | -- | 19 (4.8) |
| Doxycycline | 1 (0.3) | 1 (0.3) | -- |
| Meropenem | 1 (0.3) | -- | 29 (7.4) |
| Piperacillin-Tazobactam | -- | -- | 13 (3.3) |
| Vancomycin | -- | -- | 5 (1.3) |
| RIPE therapy | -- | -- | 5 (1.3) |
| Amikacin | -- | -- | 1 (0.3) |
| Cefepime | -- | -- | 1 (0.3) |
| Chloramphenicol | -- | -- | 1 (0.3) |
| Unknown | 9 (2.3) | -- | -- |
| Other | 6 (1.5) | -- | 15 (3.9) |

EMD, Emergency Medicine Department; RIPE, rifampin, isoniazid, pyrazinamide, ethambutol

|  | n (%) |  |  |
| --- | --- | --- | --- |
|  | Pre-Hospital | Empiric EMD | During Hospitalization |
| Any antibiotic received | 181 (46.2) | 248 (63.3) | 244 (62.2) |
| Ceftriaxone | 82 (20.9) | 228 (58.2) | 177 (45.2) |
| Metronidazole | 31 (7.9) | 100 (25.5) | 32 (8.2) |
| Amoxicillin-Clavulanate | 27 (6.9) | 13 (3.3) | 32 (8.2) |
| Gentamicin | 23 (5.9) | -- | 10 (2.6) |
| Ampicillin-Cloxacillin | 22 (5.6) | 1 (0.3) | 3 (0.8) |
| Ampicillin | 8 (2) | -- | 5 (1.3) |
| Ciprofloxacin | 7 (1.8) | 1 (0.3) | 20 (5.1) |
| Amoxicillin | 6 (1.5) | 1 (0.3) | -- |
| Penicillin | 5 (1.3) | -- | 6 (1.5) |
| Cotrimoxazole | 4 (1.0) | -- | 10 (2.6) |
| Erythromycin | 4 (1.0) | -- | -- |
| Azithromycin | 4 (1) | -- | 19 (4.8) |
| Doxycycline | 1 (0.3) | 1 (0.3) | -- |
| Meropenem | 1 (0.3) | -- | 29 (7.4) |
| Piperacillin-Tazobactam | -- | - | 13 (3.3) |
| Vancomycin | -- | -- | 5 (1.3) |
| RIPE therapy | -- | -- | 5 (1.3) |
| Amikacin | -- | -- | 1 (0.3) |
| Cefepime | -- | -- | 1 (0.3) |
| Chloramphenicol | -- | -- | 1 (0.3) |
| Unknown | 9 (2.3) | -- | -- |
| Other | 6 (1.5) | -- | 15 (3.9) |

EMD, Emergency Medicine Department; RIPE, rifampin, isoniazid, pyrazinamide, ethambutol

Baseline participant characteristics and final diagnoses between children with and without a BSI were similar (**Supplemental Tables 3 & 4)**. Among the 20 positive blood cultures, 22 pathogens were identified: 14 gram-negative bacteria (*Escherichia coli*, n = 8; *Klebsiella pneumoniae*, n = 3; *Salmonella enterica*, n = 2; *Enterobacter hormaechei*, n = 1), five gram-positive bacteria (*Streptococcus pneumoniae*, n = 4; *Staphylococcus aureus*, n = 1), and three cases of *Candida albicans* fungemia (**Figure 2A**). Mortality was highest in participants with gram-negative bacterial infections or fungemia (**Figure 2B**).

**Figure 2.**
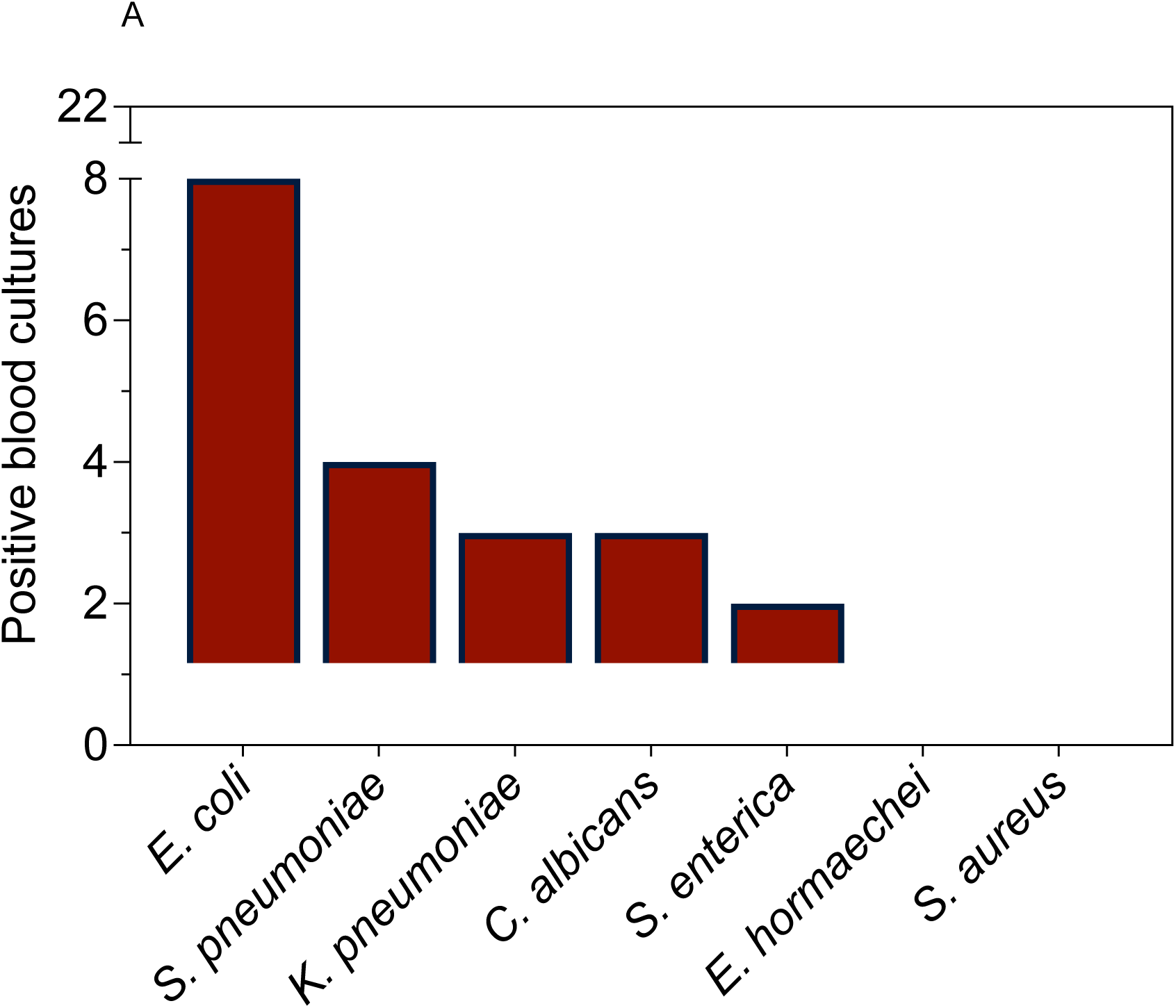
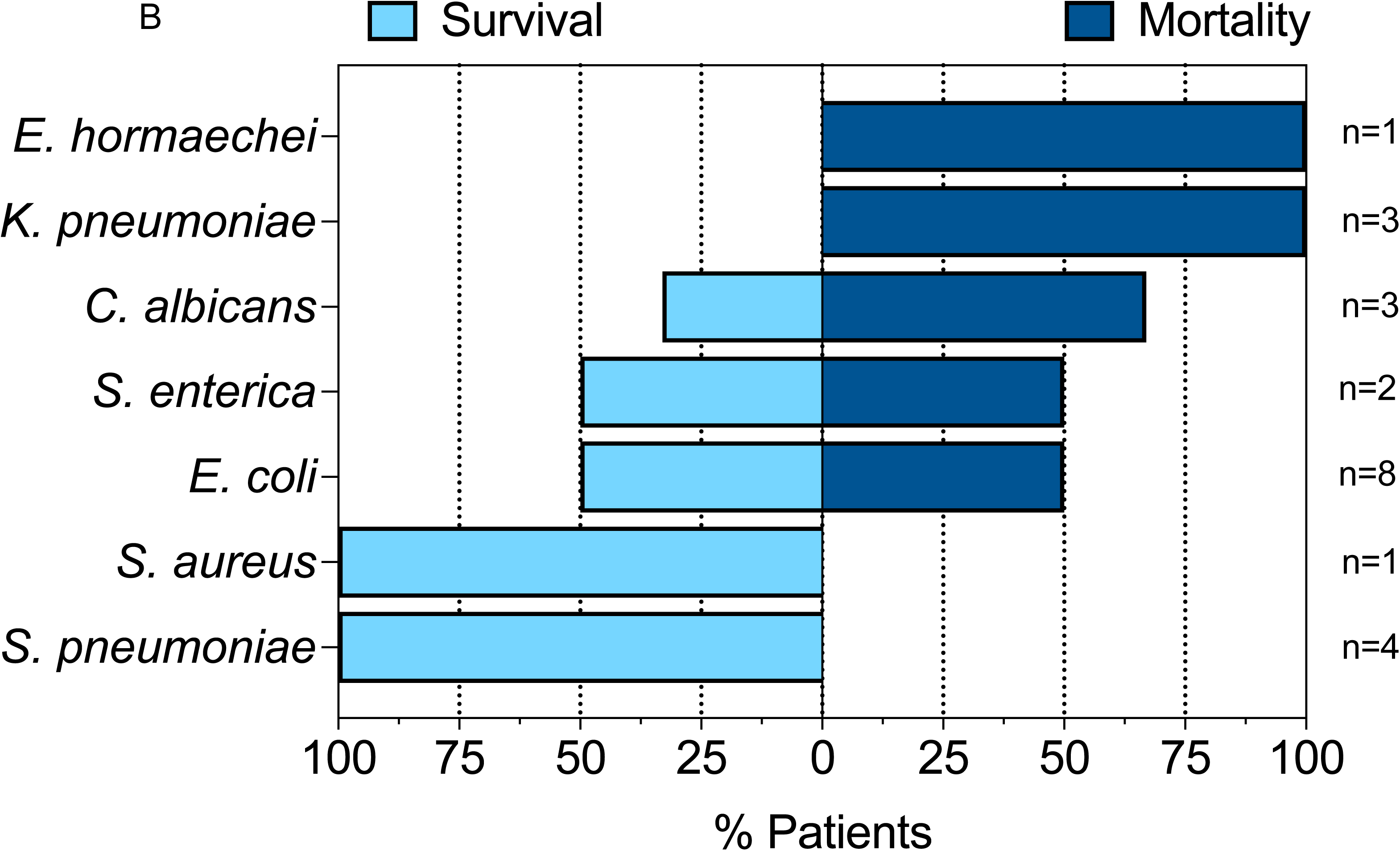
Study overview highlighting bloodstream infection (BSI) pathogens and their associations with mortality. **A)** Prevalence of BSI pathogens identified by culture. **B)** Mortality associated with BSI pathogens.

Eleven (79%) of 14 gram-negative bacterial pathogens isolated from blood culture displayed phenotypic antimicrobial susceptibility to ceftriaxone or ceftazidime resistance, suggesting ESBL production (**Figure 3A**). These 11 isolates were subsequently found to harbor *CTX-M* by WGS; *CTX-M-15* and *CTX-M-27* were detected in eight and three isolates, respectively (**Figure 4A**). Interestingly, the most common ESBL producers were *E. coli* (7 of 8 isolates, 87.5%) and *K. pneumoniae* (3 of 3 isolates, 100%). The one *E. coli* isolate that did not demonstrate an ESBL-producing phenotype had an *Amp-C* like beta-lactamase gene (*EC-5*), which can induce resistance to higher-generation cephalosporins in the presence of antibiotics.

**Figure 3.**
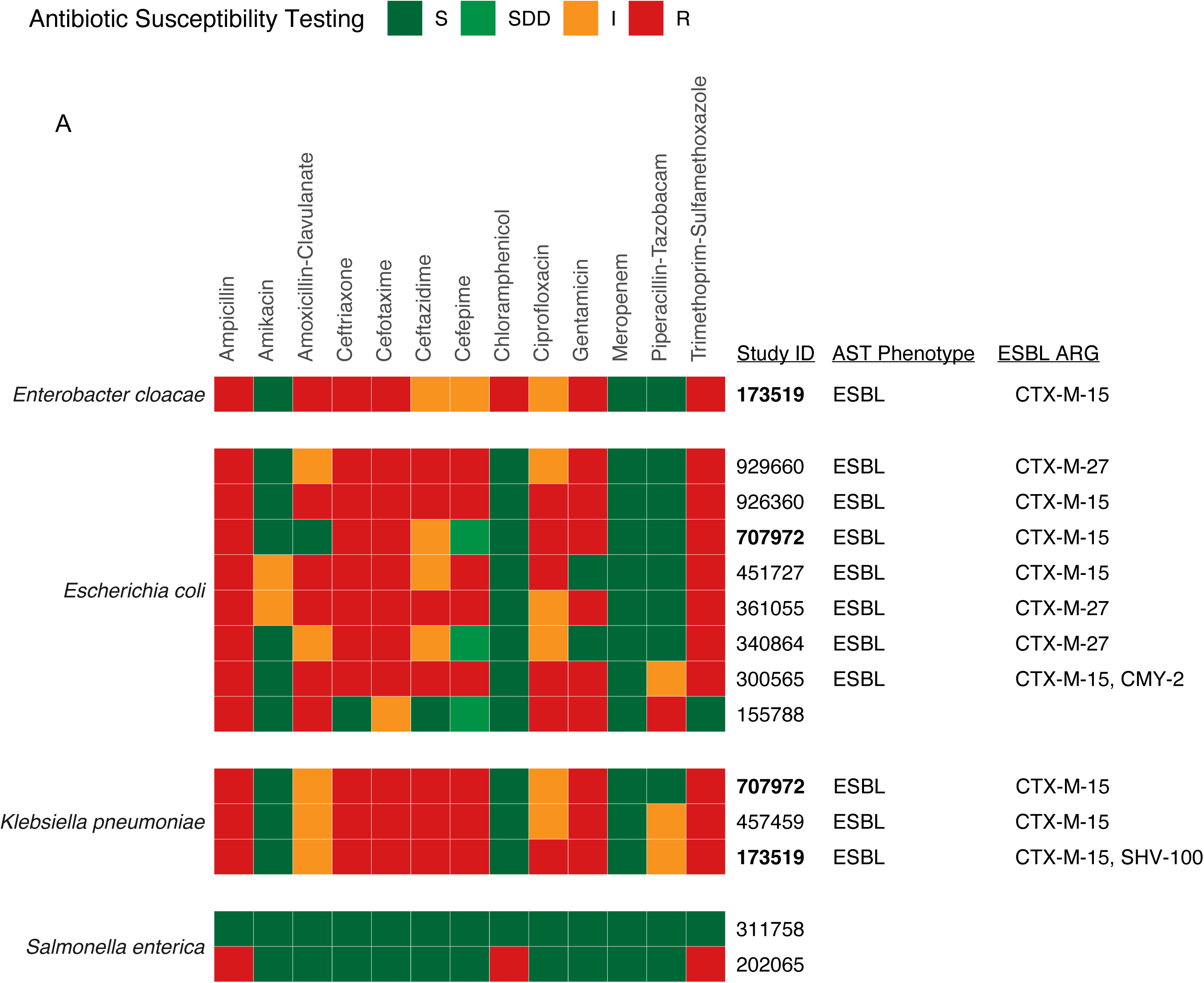
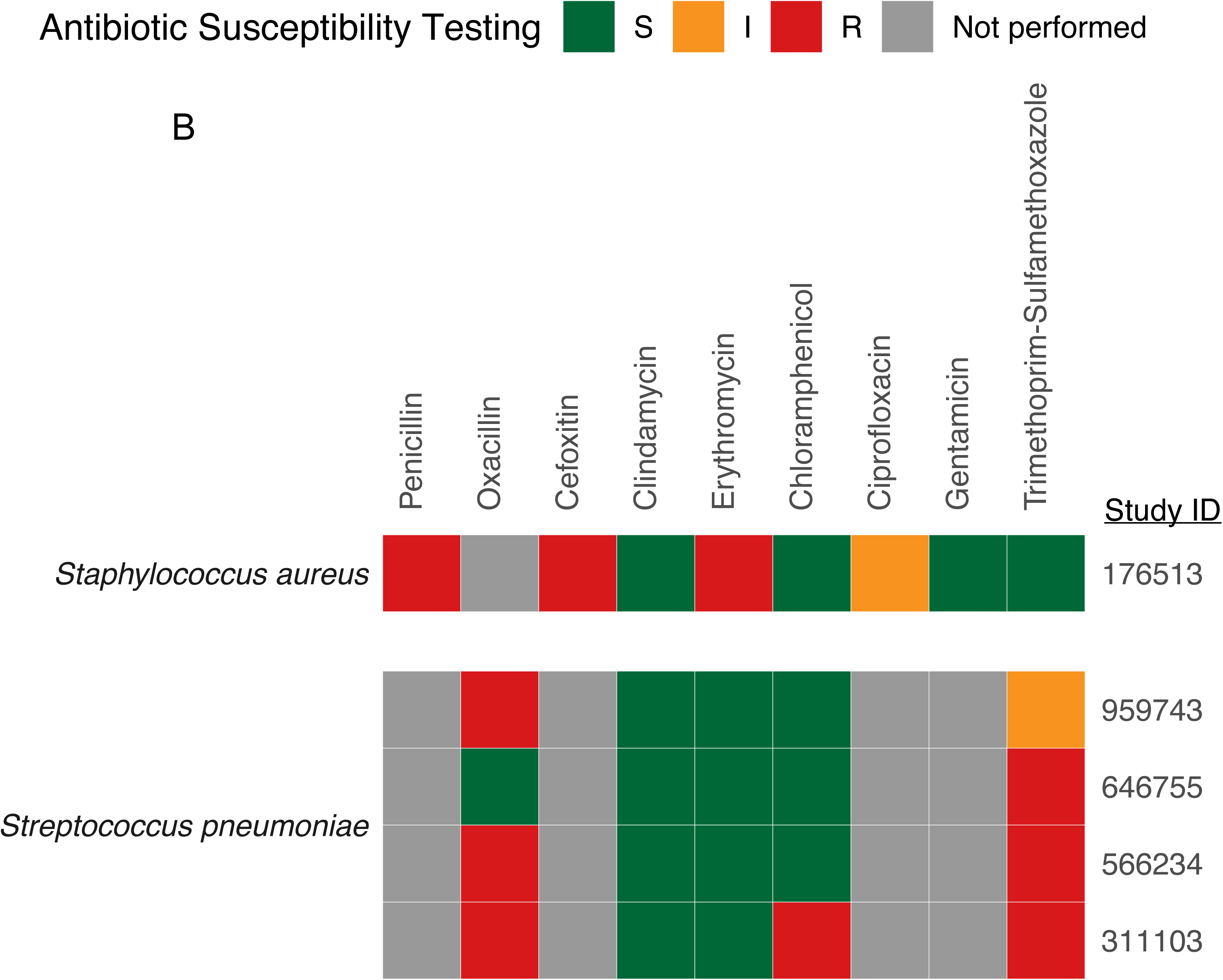
Antimicrobial resistance genes detected in bacterial blood culture isolates from Tanzanian children with severe febrile illness. Phenotypic antimicrobial sensitivity testing (AST) results and corresponding whole genome sequencing-detected antimicrobial resistance genes (ARGs) of the A) *Enterobacterales* spp. isolates and **B)** gram-positive isolates. Isolates identified by 6-digit study ID. Participants; Study IDs bolded black indicate participants with blood cultures that grew two different bacterial isolates. ESBL = extended spectrum beta lactamase. (ESBL) producers. S: Sensitive; SDD: Sensitivity Dose-Dependent; I: Intermediate; R: Resistant.

**Figure 4.**
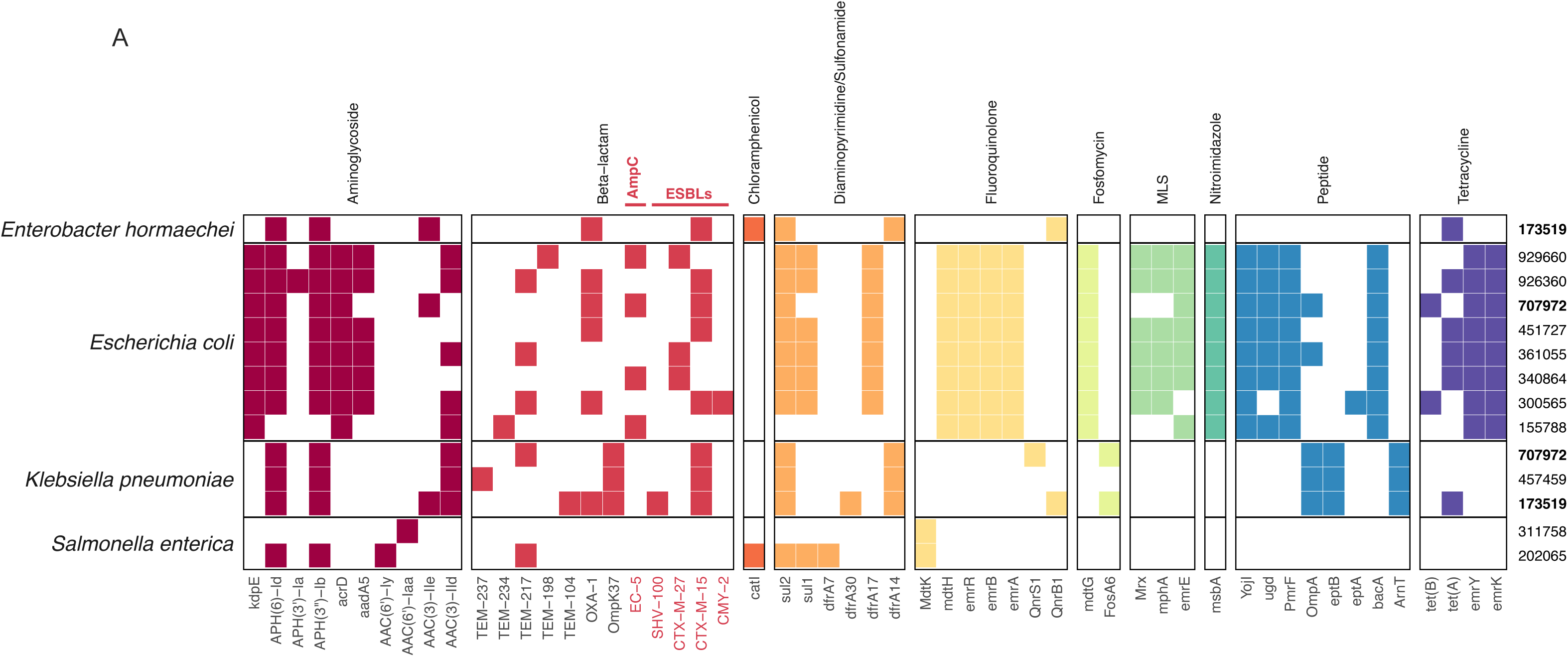
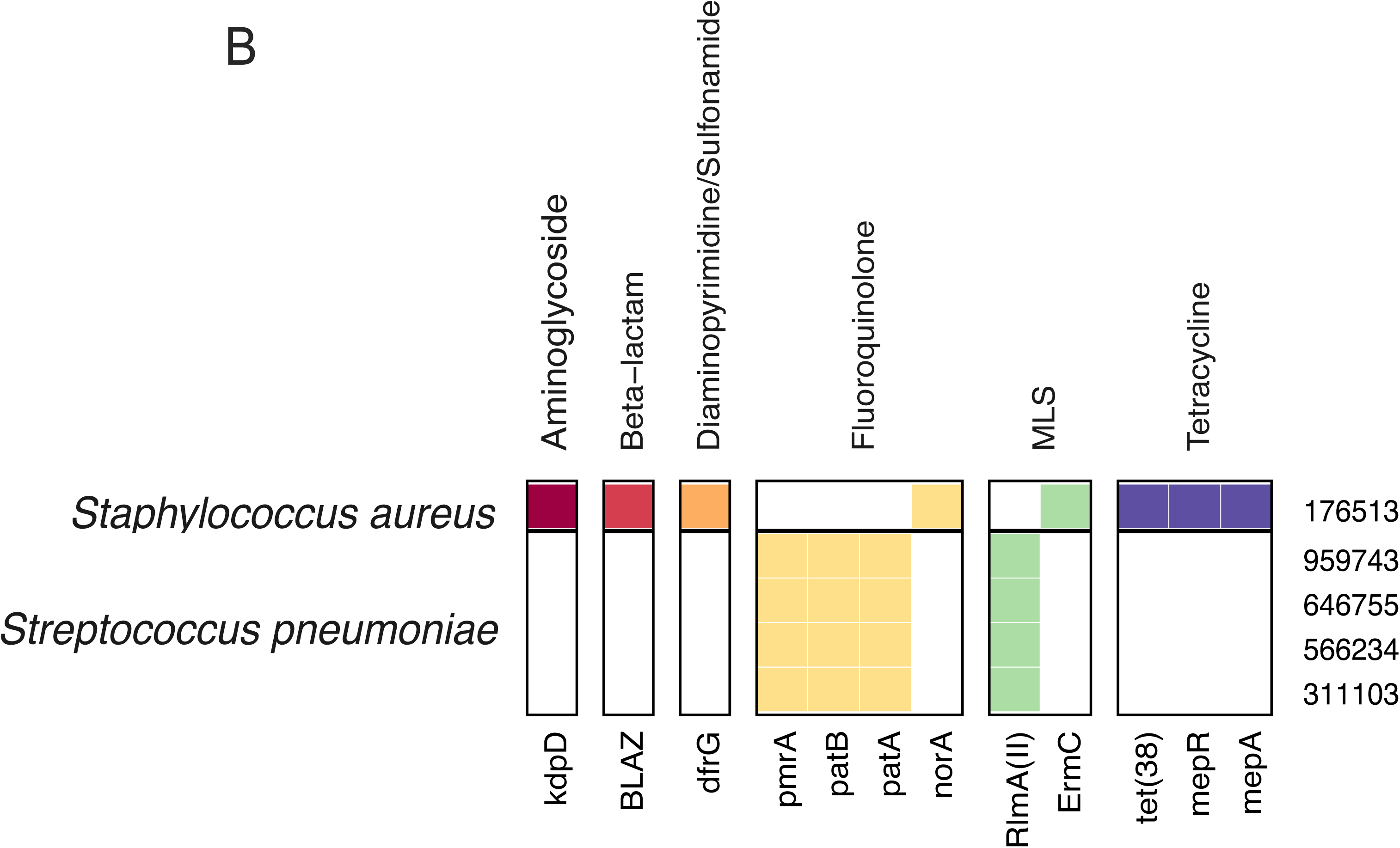
Antimicrobial resistance genes (ARGs) and genetic relatedness of bacterial bloodstream pathogens identified in children with severe febrile illness. Isolates identified by 6-digit study ID. **A)** ARGs, grouped by class, identified from WGS of gram-negative bacterial pathogens detected by blood culture. Extended-spectrum beta-lactamase and AmpC-like beta lactam resistance genes conferring ceftriaxone resistance are highlighted in red. Study IDs bolded black indicate participants with blood cultures that grew two different bacterial isolates **B)** ARGs harbored by gram-positive bacterial pathogens detected by blood culture.

Isolates from the same bacterial species shared similar ARG profiles, with the greatest intra-species variation observed in the beta-lactam ARG class (**Figure 4A**). Comparatively fewer AMR genes were identified in gram-positive BSI pathogens; the *S. aureus* isolate was methicillin-resistant (inferred from cefoxitin-resistance) and three of the four *S. pneumoniae* isolates were penicillin-resistant (inferred from oxacillin-resistance); all gram-positive isolates were susceptible to clindamycin (**Figure 3B**). Fewer ARGs were identified in gram-positive (median: 4 ARGs; IQR: 4-4 ARGs) compared with gram-negative BSI pathogens (median: 21 ARGs; IQR: 11-26 ARGs) (**Figure 4A**, **Figure 4B**).

As a secondary analysis, we sought to evaluate relationships between bacterial BSI pathogens and pneumonia pathogens isolated from tracheal aspirates of mechanically ventilated participants with clinically suspected pneumonia (n=17). Of these, seven (41.2%) had a tracheal aspirate positive for a likely pathogen including *K. pneumoniae* (n=4), *A. baumannii* (n=2), and *Pseudomonas aeruginosa* (n=1). All isolated respiratory pathogens had ESBL-producing phenotypes on susceptibility testing. From WGS, ARGs encoding ESBLs were detected in all four *K. pneumoniae* isolates (*CTX-M-15*) and both *A. baumannii* isolates (*ADC-73*) (**Supplemental Figure 1**). The four participants with *K. pneumoniae* had community-acquired infections, while those with *P. aeruginosa* and *A. baumanii* had ventilator-associated pneumonia.(36)

We next evaluated the genetic relatedness of bacterial and fungal isolates recovered from blood and tracheal aspirate cultures. None of the *E. coli* BSIs involved genetically related isolates (**Figure 5A**). Two patients were found to have identical *K. pneumoniae* infections (**Figure 5B**). Of these, participant 544023, who was hospitalized for community-acquired pneumonia, had a tracheal aspirate positive for *K. pneumoniae* that was genetically identical (0 SNPs) to an isolate from the blood of participant 707972, who had been hospitalized with sepsis over a year earlier. Two of the four patients with *S. pneumoniae* BSIs were also found to have identical isolates (**Figure 5C**).

**Figure 5.**
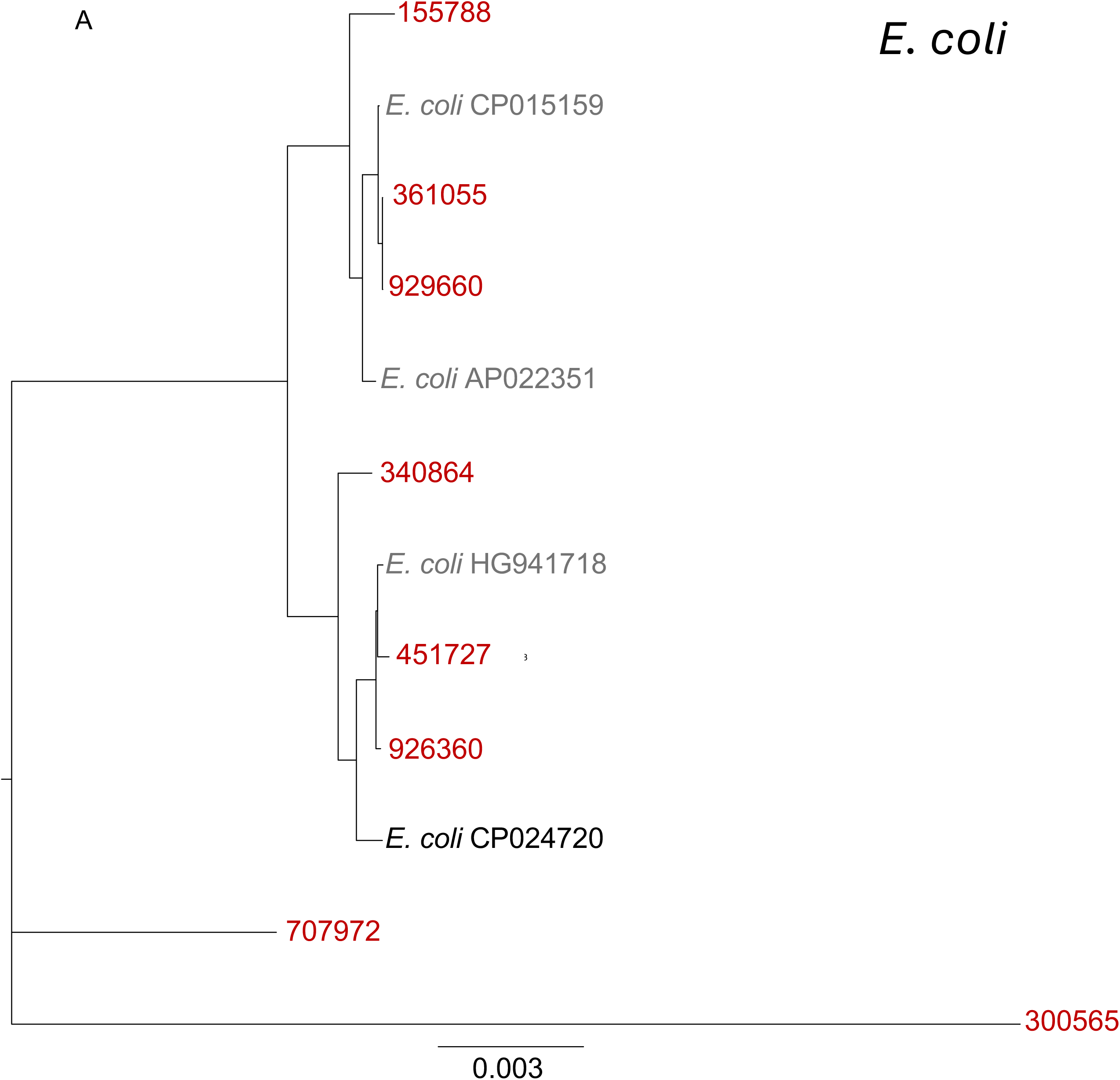
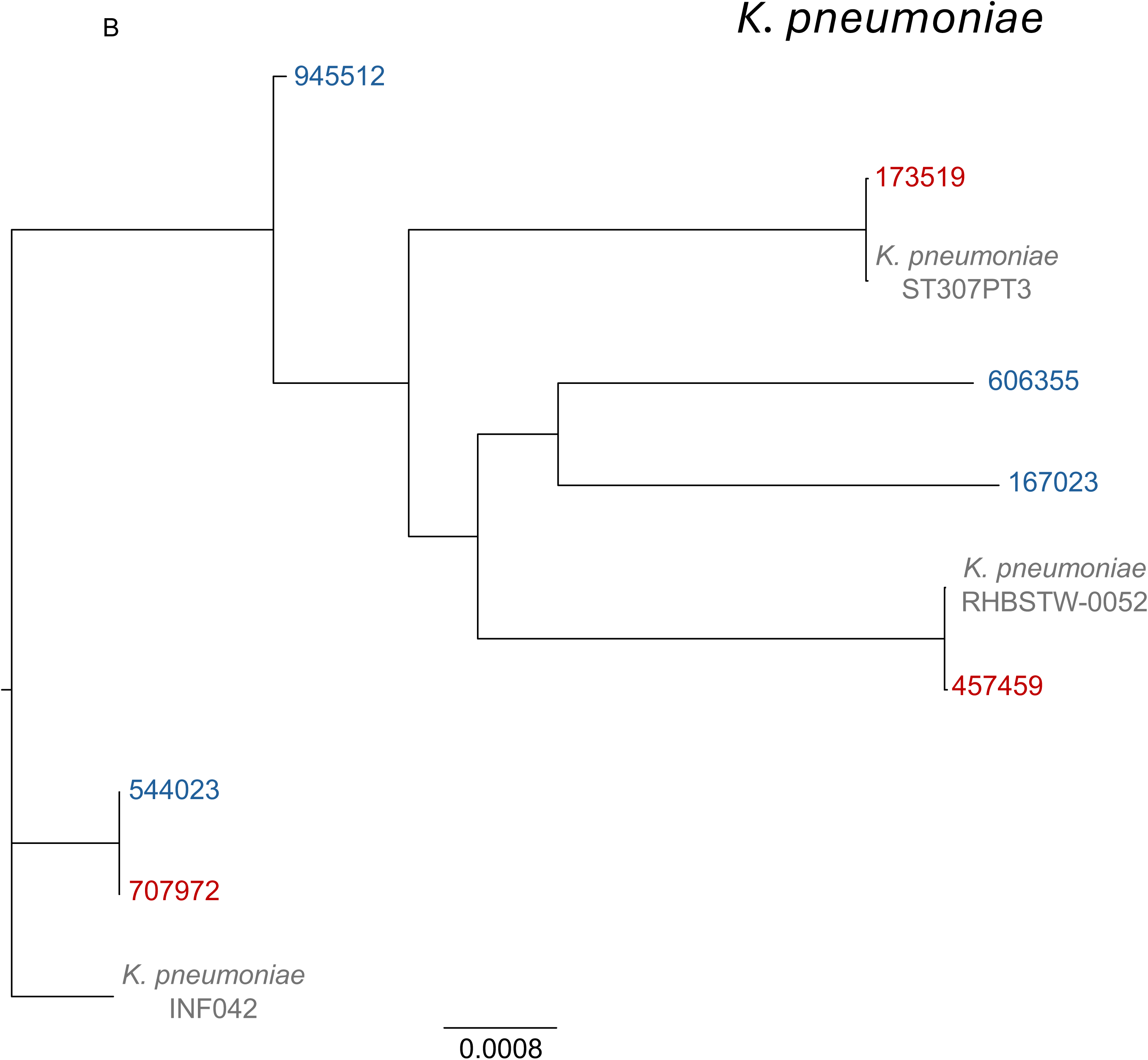
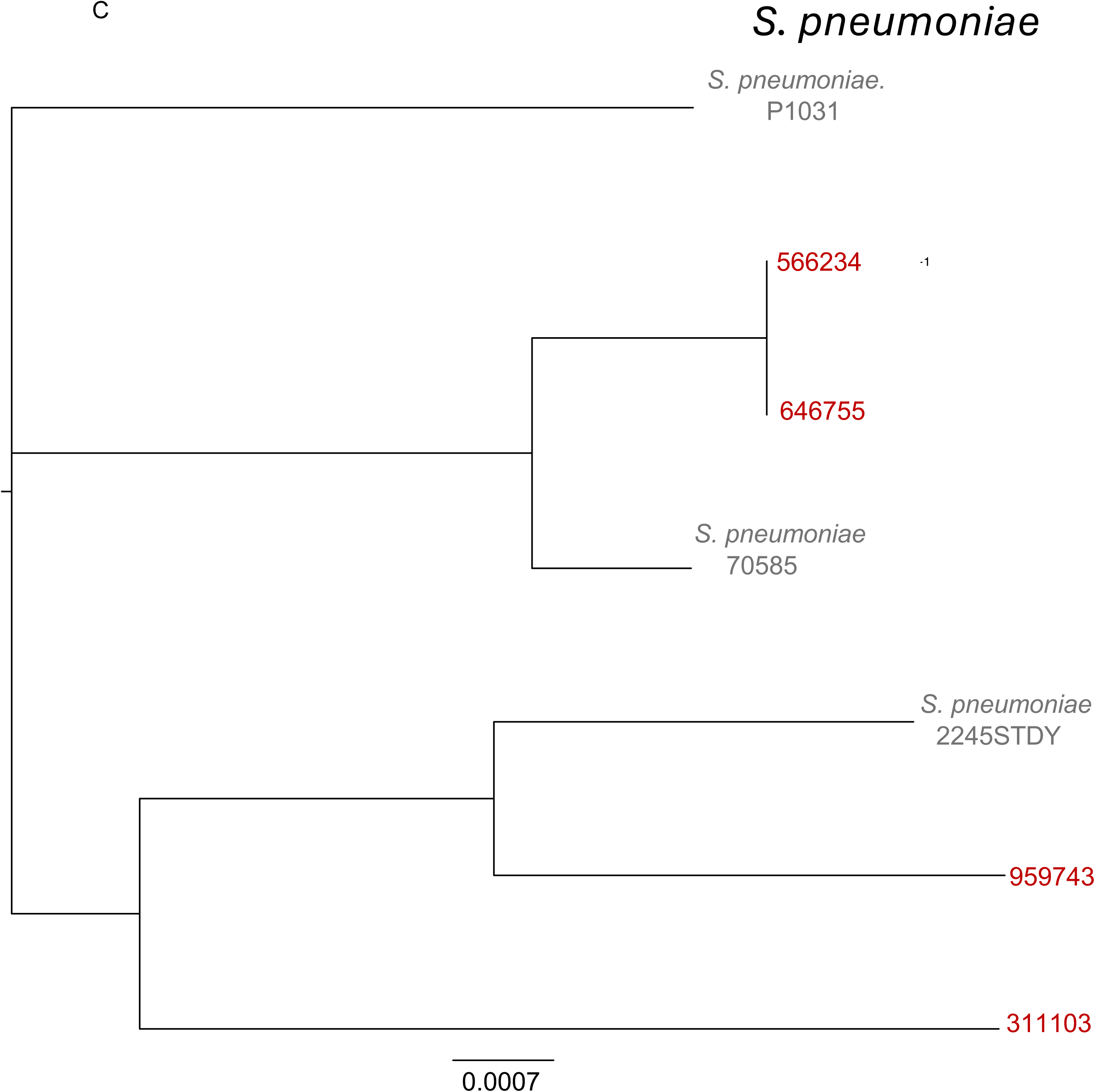
Maximum likelihood phylogenetic trees for blood culture isolates (subject IDs depicted in red) and tracheal aspirate cultures (subject IDs depicted in blue) for **A)** *E. coli* isolates demonstrating that community-onset bloodstream infections from this pathogen were unrelated in this cohort. **B)** *K. pneumoniae* isolates demonstrating that community-onset bloodstream infections were unrelated but that a tracheal aspirate isolate was genetically related to a BSI from a patient hospitalized over one year earlier. **C)** *S. pneumoniae* isolates demonstrating shared community-onset infections between two patients. MLS: macrolide-lincosamide-streptogramin. Red = blood culture isolates. Blue = tracheal aspirate isolates. Grey = reference genomes. Scale bar indications substitutions per site.

In addition, two of the three participants with candidemia had genetically identical *C. albicans* strains recovered from their blood cultures collected at presentation (**Supplemental Figure 2**). The two participants with positive tracheal aspirate cultures for *A. baumanii* had genetically related isolates (1 SNP difference, **Supplemental Figure 3**), demonstrating that two of seven (28.6%) tracheal aspirate cultures represented probable nosocomial infections.

## Discussion

In a prospective cohort of 392 Tanzanian children with SFI, we found that culture-confirmed BSI is associated with a four-fold increase in odds of mortality. Furthermore, we reported that the majority of identified BSI pathogens are resistant to ceftriaxone, the first-line antimicrobial regimen recommended by National Guidelines for the treatment of SFI.(37) Using WGS, we further demonstrated that the majority of gram-negative pathogens harbored *CTX-M* or other ESBL genes, and uncovered evidence of pathogen transmission in both the community and hospital.

Mortality in participants with BSI was 45%, notably higher than the 8% mortality observed in a previous cohort of young children with acute febrile illness hospitalized in four Tanzanian public hospitals, including MNH.(4) Mortality in our cohort was also higher than that in a study of 19 sites primarily in LMICs, which reported 18% mortality in infants hospitalized with sepsis due to a pathogen-positive culture.(38) These differences may be due to delayed presentation and/or increased severity of illness at our study site, a national referral hospital.(39)

The prevalence of BSI (5%) in our cohort was lower than in a previous Tanzanian pediatric acute febrile illness cohort (11%) or a global observational study of infants with sepsis (18%).(4, 38) As our study site was a national referral hospital, participants were frequently evaluated at other facilities prior to presentation, which presumably contributed to the high rate of pre-hospital antibiotics (46%), which may have rendered cultures falsely negative.

Using locally available clinical diagnostics, a definitive BSI pathogen (bacterial, fungal, or parasite) was identified in only 10% of participants, emphasizing that the underlying infectious etiology of SFI is not identified in most cases. In a large systematic review of SFI in East Africa (29,286 adults and children) that included studies incorporating more comprehensive diagnostics (e.g., PCR, serology), pathogens were identified in only 18% of participants.(8) Together, this emphasizes the urgent need for more advanced, culture-independent diagnostic approaches such as metagenomic sequencing to improve our understanding of SFI etiologies in this population.

Importantly, among identified pathogens, phenotypic and genotypic AMR analyses demonstrated a high prevalence of resistance among *Enterobacterales* to ceftriaxone, which is currently the first-line empiric antibiotic for both outpatient and inpatient pediatric febrile illness at MNH. Prior studies in pediatric patients in Tanzania and elsewhere in SSA demonstrate a high frequency of ceftriaxone use for empiric management of infections, primarily in hospitalized patients, which is consistent with both local and national guidelines.(37, 40–43) Ceftriaxone is widely available, relatively low-cost, and thought to provide adequate coverage for pathogens associated with common pediatric bacterial infections. Its primary coverage gaps include ESBL and *AmpC*-producing gram-negative organisms, anaerobes, and some gram-positive pathogens such as *Staphylococcus* or *Enterococcus* spp.

Among the identified BSI pathogens, ceftriaxone would normally be considered appropriate coverage for 18 of the 22 pathogens detected (excluding *S. aureus* bacteremia or fungemia), and its empiric use would have been effective. However, due to the high prevalence of ESBL-production among the *Enterobacterales* pathogens detected, only seven pathogens (4/4 *S. pneumoniae*, 2/2 *S. enterica*, and 1/8 *E. coli*) were ceftriaxone-susceptible. Furthermore, all ESBL-producing blood pathogens had *CTX-M*, the most common ESBL gene type worldwide.(44) This aligns with findings from other pediatric AMR studies in SSA that demonstrate a high prevalence of ESBL-producing organisms, including a recent cross-sectional study of young children in Tanzania where over 50% of hospitalized children with bacteremia were infected with ESBL and/or multi-drug resistant organisms.(4, 9, 45)

The lack of genetic relatedness between *E. coli* isolates reinforces the significance and pervasiveness of AMR in the community. The phylogenetic analyses of the *K. pneumoniae* isolates, however, suggested transmission of pathogenic strains in the community. We detected identical *K. pneumoniae* isolates from two separate participants enrolled over a year apart, one with culture-confirmed community-acquired BSI and the second with community-acquired pneumonia, suggesting sustained circulation of the ESBL-producing *K. pneumoniae* strain in the community. Concerningly, multiple studies demonstrate an increased prevalence of ESBL-producing organisms circulating in the community and causing community-acquired infections.(46–48)

In our secondary analysis of tracheal aspirate cultures from mechanically ventilated patients, we found evidence of nosocomial transmission in two cases of *A. baumanii* ventilator-associated pneumonia. *A. baumanii* is a problematic and frequently multi-drug-resistant nosocomial pathogen,(49) and these findings emphasize that strong infection prevention measures are imperative to prevent hospital-acquired infections, especially among the most vulnerable patient populations. Additionally, the integration of clinical and genomic data is a powerful tool for understanding and, ultimately, preventing nosocomial transmission. Further work is needed to understand potential transmission sources (e.g., staff, medical equipment, other environmental factors) for these infections.

This study has several strengths; it is a prospective pediatric cohort in SSA where the etiology of SFI is not well understood, it includes detailed clinical phenotyping, and incorporates WGS. It also has limitations, including a reliance on bacterial and fungal cultures for pathogen detection. Although microbial culture is the gold standard, culture-independent techniques (e.g., metagenomic sequencing) may have identified additional missed infections, including those pre-treated with antibiotics. Second, the study site was a tertiary care referral hospital that cares for the most severe cases and the cohort may not be representative of the population cared for in lower-level hospitals. Third, the relatively small number of bacterial and fungal pathogens isolated in culture limited our capacity to rigorously evaluate pathogen-specific associations with mortality. Fourth, not all participants with suspected pneumonia were intubated, and tracheal aspirate collection was determined by the treating clinician; accordingly, our results likely underestimate the incidence of severe bacterial pneumonia and nosocomial transmission.

In conclusion, among children with SFI, BSI conferred high mortality and was frequently caused by ESBL-producing gram-negative organisms resistant to first-line antibiotics. Additional surveillance studies across SSA are needed to confirm these findings and, if warranted, to guide revision of empiric antimicrobial regimens. Reducing the burden of pediatric SFI and AMR requires a multifaceted approach including prevention, evidenced-based empiric antimicrobial use, improving access to timely and accurate diagnostics, and increasing AMR surveillance.(13, 50)

## Acknowledgments

We would like to thank Amy Markowitz for a thoughtful review of the manuscript draft.

## About the Author

Teresa Kortz is an Associate Professor of Clinical Pediatrics in Critical Care Medicine at the University of California, San Francisco. Dr. Kortz has a PhD in Global Health Sciences and is an NIH-funded researcher who studies pediatric severe febrile illness etiology, prognostic biomarkers, and clinical outcomes in East Africa.

## Disclaimer

The opinions expressed by authors contributing to this journal do not necessarily reflect the opinions of the Centers for Disease Control and Prevention or the institutions with which the authors are affiliated.

## Data Availability

Raw sequencing reads are available from the NCBI Sequence Read Archive accession SUB14909672.

## Supplemental Tables

**Supplemental Table 1.** Strobe checklist for observational cohort studies

**Supplemental Table 2.**
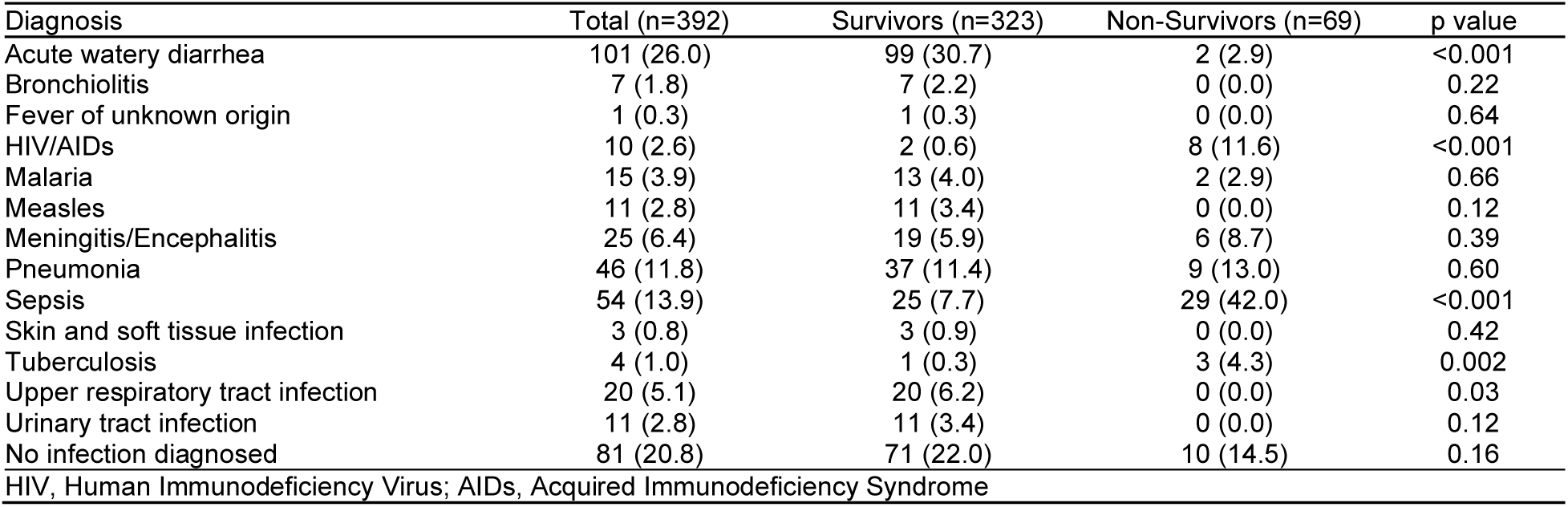
Final infectious diagnosis at time of hospital discharge or death for children with severe febrile illness in Tanzania by survival status.

**Supplemental Table 3.**
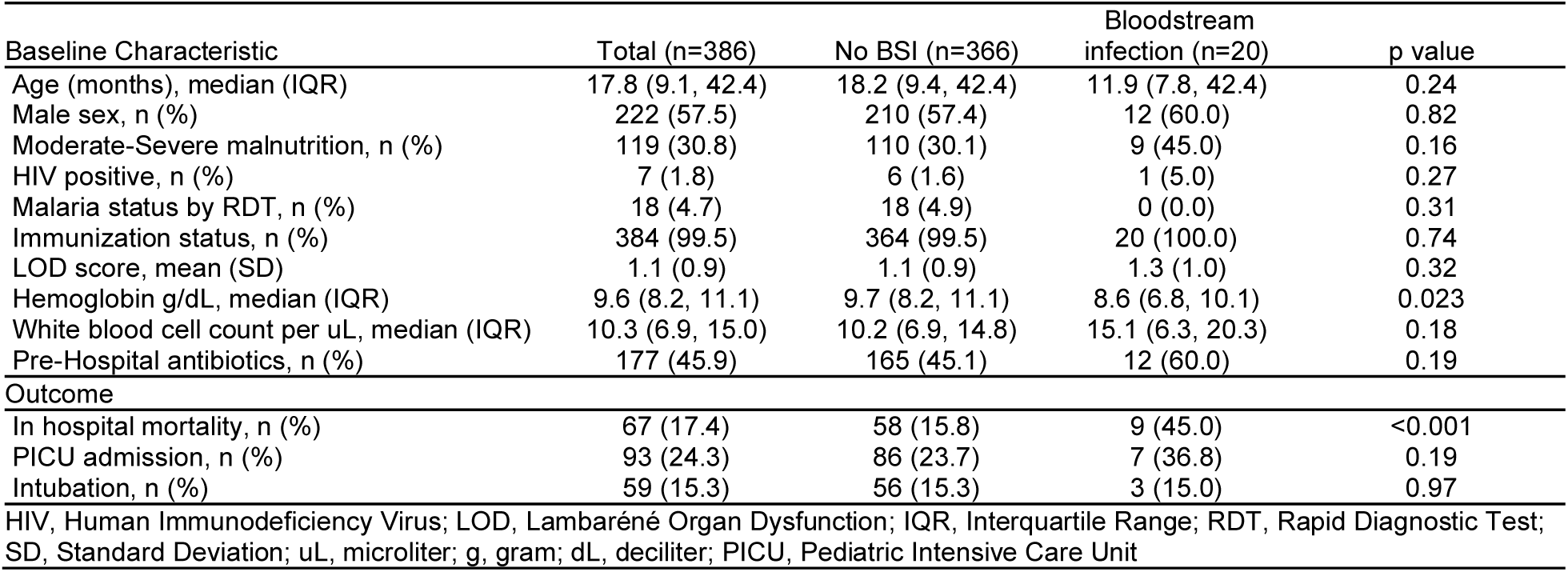
Participant baseline characteristics and outcomes for children with severe febrile illness in Tanzania by bacteremia status; limited to those with blood culture results (n=386).

**Supplemental Table 4.**
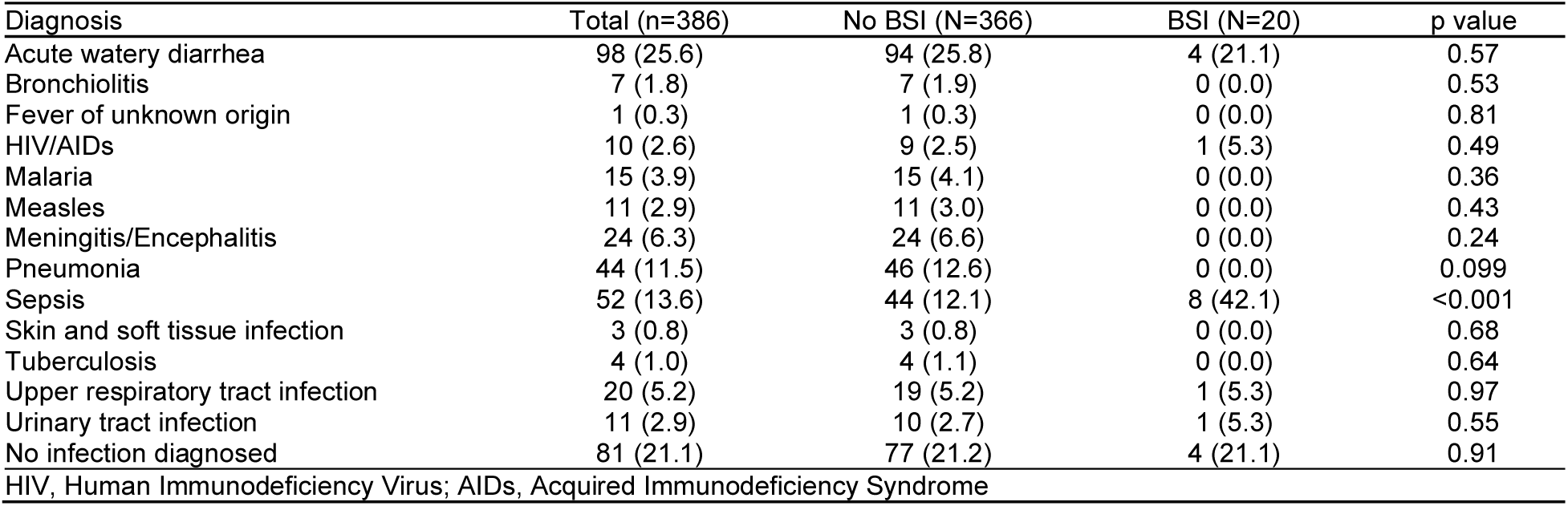
Final infectious diagnosis at time of hospital discharge or death for children with severe febrile illness in Tanzania by bacteremia status; limited to those with blood culture results (n=386).

## Supplemental Figures

**Supplemental Figure 1. Antimicrobial resistance of gram-negative bacterial isolates from tracheal aspirate cultures of Tanzanian children with severe febrile illness.** A) Phenotypic antimicrobial sensitivity testing (AST) results and whole genome sequencing-detected antimicrobial resistance genes (ARGs) of gram-negative isolates. B) All detected ARGs, grouped by ARG class, for each isolate. Participants with ceftriaxone-resistant isolates are extended-spectrum beta-lactamase (ESBL) producers, and those with meropenem resistance are carbapenem-resistant organisms (CRO). Isolates identified by 6-digit study ID. S: Sensitive; SDD: Sensitivity Dose-Dependent; I: Intermediate; R: Resistant; MLS: macrolide-lincosamide-streptogramin.

**Supplemental Figure 2. Phylogenetic analysis of *C. albicans* blood culture isolates.** Maximum likelihood phylogenetic tree. Isolates identified in red by 6-digit study ID, reference genome in grey. Scale bar indications substitutions per site.

**Supplemental Figure 3. Phylogenetic analysis of *A. baumanii* isolates from tracheal aspirate cultures.** Maximum likelihood phylogenetic tree demonstrating probable nosocomial transmission of *A. baumanii* in two patients with ventilator-associated pneumonia. Isolates identified in blue by 6-digit study ID, reference genomes in grey. Scale bar indications substitutions per site.

