## Supplementary figures and images for "Prospective Genomic Surveillance of Severe Febrile Illness in Tanzanian Children Identifies High Mortality and Resistance to First-Line Antibiotics in Bloodstream Infections"

### Supplemental Figure 1A

Supplemental Figure 1A

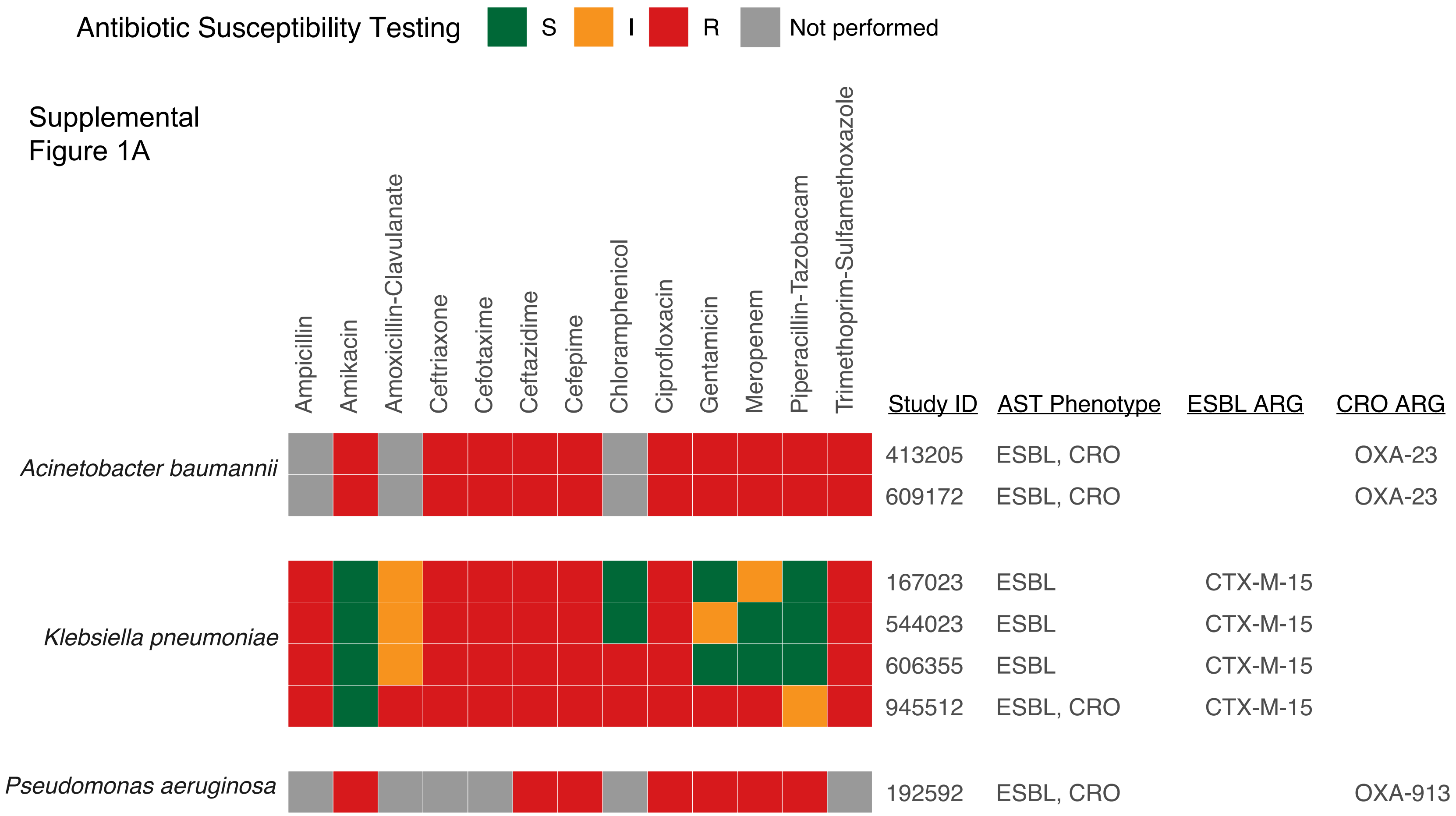

### Supplemental Figure 1B

Supplemental Figure 1B

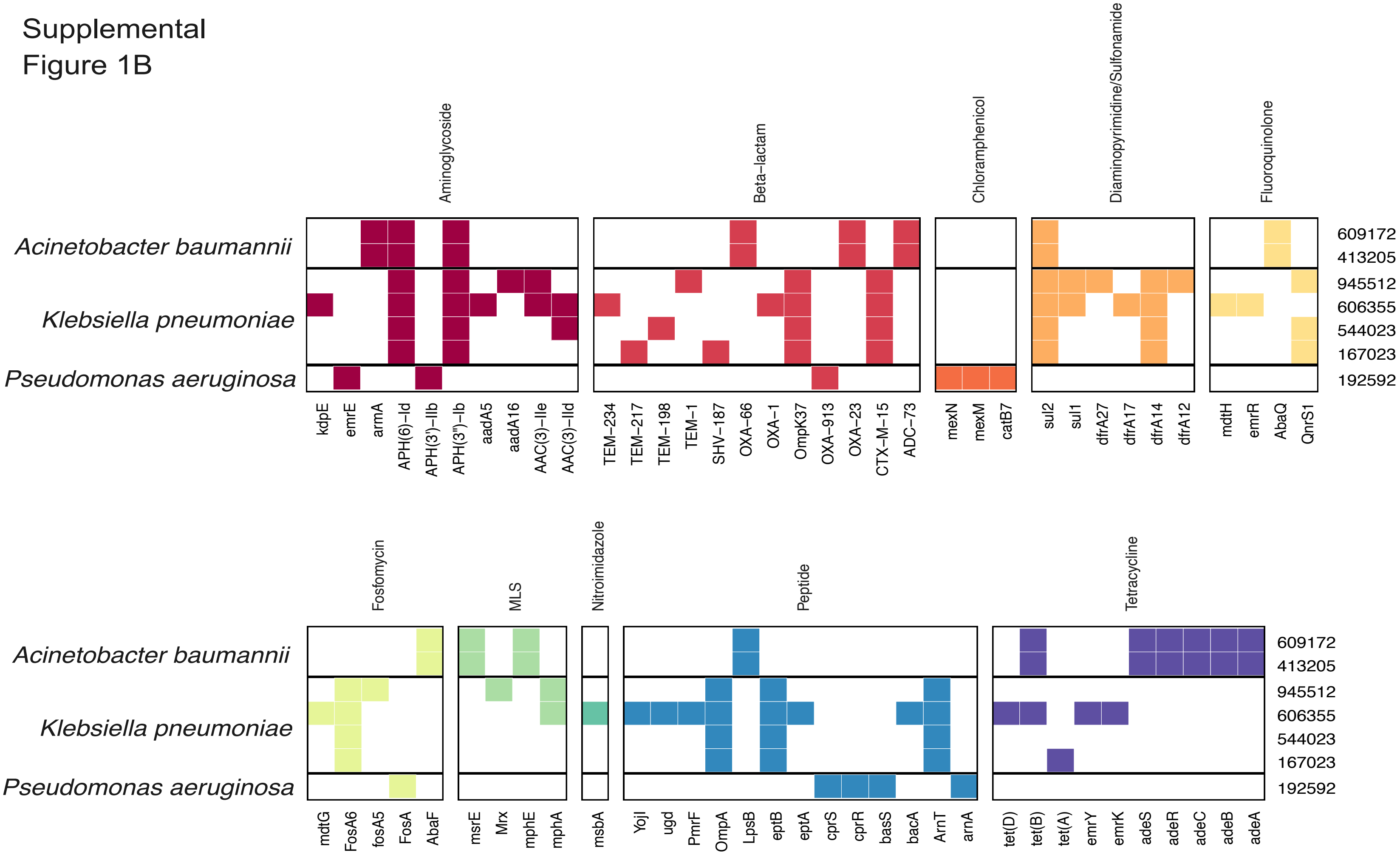

### Supplemental Figure 2

Supplemental Figure 2

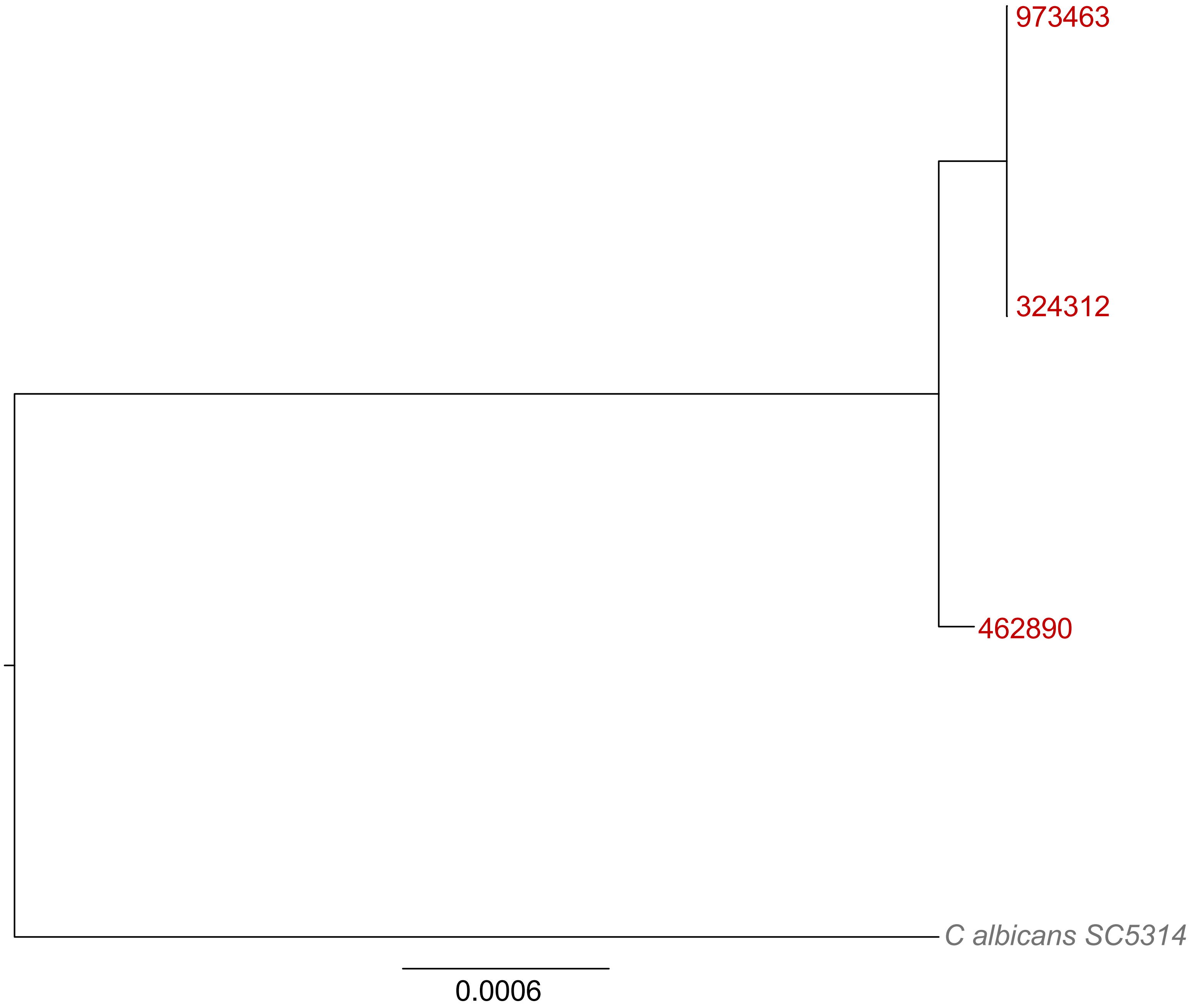

### Supplemental Figure 3

Supplemental Figure 3

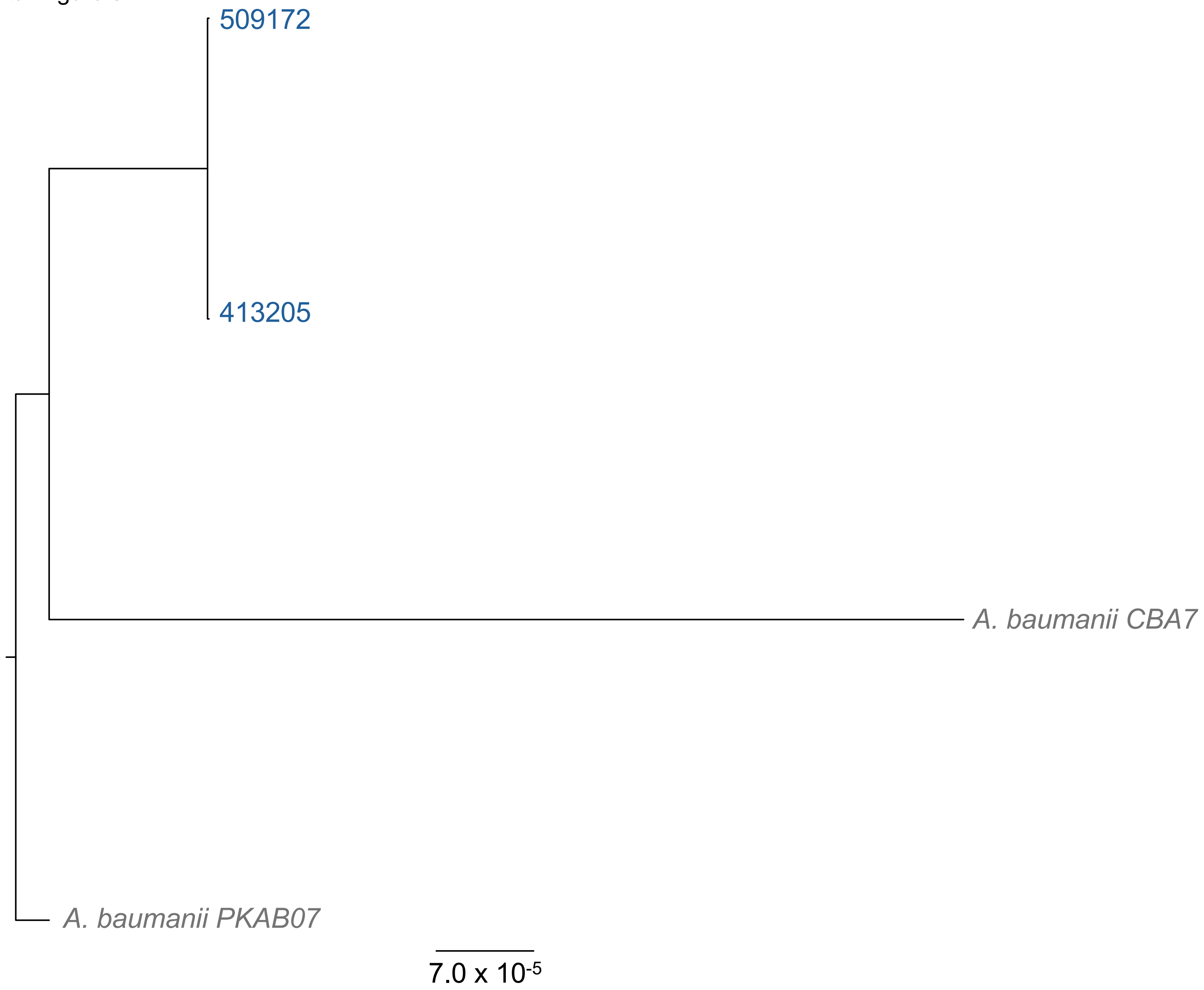
