## Supplemental Table 2 for "Prospective Genomic Surveillance of Severe Febrile Illness in Tanzanian Children Identifies High Mortality and Resistance to First-Line Antibiotics in Bloodstream Infections"

**Supplemental Table 2**. Final infectious diagnosis at time of hospital discharge or death for children with severe febrile illness in Tanzania by survival status

| Diagnosis | Total (n=392) | Survivors (n=323) | Non-Survivors (n=69) | p value |
| --- | --- | --- | --- | --- |
| Acute watery diarrhea | 101 (26.0) | 99 (30.7) | 2 (2.9) | <0.001 |
| Bronchiolitis | 7 (1.8) | 7 (2.2) | 0 (0.0) | 0.22 |
| Fever of unknown origin | 1 (0.3) | 1 (0.3) | 0 (0.0) | 0.64 |
| HIV/AIDs | 10 (2.6) | 2 (0.6) | 8 (11.6) | <0.001 |
| Malaria | 15 (3.9) | 13 (4.0) | 2 (2.9) | 0.66 |
| Measles | 11 (2.8) | 11 (3.4) | 0 (0.0) | 0.12 |
| Meningitis/Encephalitis | 25 (6.4) | 19 (5.9) | 6 (8.7) | 0.39 |
| Pneumonia | 46 (11.8) | 37 (11.4) | 9 (13.0) | 0.60 |
| Sepsis | 54 (13.9) | 25 (7.7) | 29 (42.0) | <0.001 |
| Skin and soft tissue infection | 3 (0.8) | 3 (0.9) | 0 (0.0) | 0.42 |
| Tuberculosis | 4 (1.0) | 1 (0.3) | 3 (4.3) | 0.002 |
| Upper respiratory tract infection | 20 (5.1) | 20 (6.2) | 0 (0.0) | 0.03 |
| Urinary tract infection | 11 (2.8) | 11 (3.4) | 0 (0.0) | 0.12 |
| No infection diagnosed | 81 (20.8) | 71 (22.0) | 10 (14.5) | 0.16 |
| HIV, Human Immunodeficiency Virus; AIDs, Acquired Immunodeficiency Syndrome | | | | |
