## Supplemental Table 3 for "Prospective Genomic Surveillance of Severe Febrile Illness in Tanzanian Children Identifies High Mortality and Resistance to First-Line Antibiotics in Bloodstream Infections"

**Supplemental Table 3.** Participant baseline characteristics and outcomes for children with severe febrile illness in Tanzania by bloodstream infection (BSI) status; limited to those with blood culture results (n=386)

| Baseline Characteristic | Total (n=386) | No BSI (n=366) | Bloodstream infection (n=20) | p value |
| --- | --- | --- | --- | --- |
| Age (months), median (IQR) | 17.8 (9.1, 42.4) | 18.2 (9.4, 42.4) | 11.9 (7.8, 42.4) | 0.24 |
| Male sex, n (%) | 222 (57.5) | 210 (57.4) | 12 (60.0) | 0.82 |
| Moderate-Severe malnutrition, n (%) | 119 (30.8) | 110 (30.1) | 9 (45.0) | 0.16 |
| HIV positive, n (%) | 7 (1.8) | 6 (1.6) | 1 (5.0) | 0.27 |
| Malaria status by RDT, n (%) | 18 (4.7) | 18 (4.9) | 0 (0.0) | 0.31 |
| Immunization status, n (%) | 384 (99.5) | 364 (99.5) | 20 (100.0) | 0.74 |
| LOD score, mean (SD) | 1.1 (0.9) | 1.1 (0.9) | 1.3 (1.0) | 0.32 |
| Hemoglobin g/dL, median (IQR) | 9.6 (8.2, 11.1) | 9.7 (8.2, 11.1) | 8.6 (6.8, 10.1) | 0.023 |
| White blood cell count per uL, median (IQR) | 10.3 (6.9, 15.0) | 10.2 (6.9, 14.8) | 15.1 (6.3, 20.3) | 0.18 |
| Pre-Hospital antibiotics, n (%) | 177 (45.9) | 165 (45.1) | 12 (60.0) | 0.19 |
| Outcome |  |  |  |  |
| In hospital mortality, n (%) | 67 (17.4) | 58 (15.8) | 9 (45.0) | <0.001 |
| PICU admission, n (%) | 93 (24.3) | 86 (23.7) | 7 (36.8) | 0.19 |
| Intubation, n (%) | 59 (15.3) | 56 (15.3) | 3 (15.0) | 0.97 |
| HIV, Human Immunodeficiency Virus; LOD, Lambaréné Organ Dysfunction; IQR, Interquartile Range; RDT, Rapid Diagnostic Test; SD, Standard Deviation; uL, microliter; g, gram; dL, deciliter; PICU, Pediatric Intensive Care Unit | | | | |
