## Supplemental Table 4 for "Prospective Genomic Surveillance of Severe Febrile Illness in Tanzanian Children Identifies High Mortality and Resistance to First-Line Antibiotics in Bloodstream Infections"

**Supplemental Table 4**. Final infectious diagnosis at time of hospital discharge or death for children with severe febrile illness in Tanzania by bloodstream infection (BSI) status; limited to those with blood culture results (n=386)

| Diagnosis | Total (n=386) | No BSI (N=366) | BSI (N=20) | p value |
| --- | --- | --- | --- | --- |
| Acute watery diarrhea | 98 (25.6) | 94 (25.8) | 4 (21.1) | 0.57 |
| Bronchiolitis | 7 (1.8) | 7 (1.9) | 0 (0.0) | 0.53 |
| Fever of unknown origin | 1 (0.3) | 1 (0.3) | 0 (0.0) | 0.81 |
| HIV/AIDs | 10 (2.6) | 9 (2.5) | 1 (5.3) | 0.49 |
| Malaria | 15 (3.9) | 15 (4.1) | 0 (0.0) | 0.36 |
| Measles | 11 (2.9) | 11 (3.0) | 0 (0.0) | 0.43 |
| Meningitis/Encephalitis | 24 (6.3) | 24 (6.6) | 0 (0.0) | 0.24 |
| Pneumonia | 44 (11.5) | 46 (12.6) | 0 (0.0) | 0.099 |
| Sepsis | 52 (13.6) | 44 (12.1) | 8 (42.1) | <0.001 |
| Skin and soft tissue infection | 3 (0.8) | 3 (0.8) | 0 (0.0) | 0.68 |
| Tuberculosis | 4 (1.0) | 4 (1.1) | 0 (0.0) | 0.64 |
| Upper respiratory tract infection | 20 (5.2) | 19 (5.2) | 1 (5.3) | 0.97 |
| Urinary tract infection | 11 (2.9) | 10 (2.7) | 1 (5.3) | 0.55 |
| No infection diagnosed | 81 (21.1) | 77 (21.2) | 4 (21.1) | 0.91 |
| HIV, Human Immunodeficiency Virus; AIDs, Acquired Immunodeficiency Syndrome | | | | |
